# Pharmacists’ and Pharmacy Technicians’ Attitudes and Experiences with Technician Administered Immunizations

**DOI:** 10.1101/2022.06.10.22276245

**Authors:** Alexis DiMario, Kenneth L McCall, Sara Couture, Wendy Boynton

**Affiliations:** School of Pharmacy, University of New England, Portland, Maine, USA; Hannaford Supermarkets & Pharmacy, Scarborough, Maine, USA; Binghamton University School of Pharmacy & Pharmaceutical Sciences, Johnson City, NY, USA

## Abstract

**Background:** In response to the increased demand for healthcare services during the COVID-19 pandemic, the Public Readiness and Emergency Preparedness (PREP) Act amendments and guidance authorized pharmacy technicians, who are not otherwise authorized in their state, to administer the Advisory Committee on Immunization Practices (ACIP) recommended immunizations and COVID-19 vaccines under pharmacist order. Subsequently, many pharmacies nationwide have expanded technician duties to include immunization administration.

**Objectives:** The primary objective of this study is to evaluate and compare the attitudes and experiences associated with technician administered immunizations among community pharmacists and technicians.

**Methods:** The cross-sectional study evaluated the primary endpoint through the completion of anonymous surveys containing peer reviewed questionnaires. Pharmacy technicians and their supervising pharmacists were selected to complete the survey at a grocery chain pharmacy located in 5 states across the Northeast if they completed the APhA immunization program and administered at least 1 immunization. Surveys were drafted using Microsoft Forms and results were analyzed using Microsoft Excel. Chi square tests were utilized to evaluate the correlation between responses.

**Results:** A total of 268 survey responses was obtained with 171 responses coming from pharmacists and 97 responses coming from immunization certified technicians. Most pharmacists and pharmacy technicians responded that technicians could safely administer vaccines (87.1% and 96.9%, respectively) and competently process and bill vaccine services (90.6% and 99.0%, respectively). In addition, both participant populations responded that technician administered vaccines improved the workflow of vaccine services (76.6% and 82.5%, respectively) without increasing the likelihood of vaccine errors (56.1% and 78.3%, respectively). When compared with technicians, fewer pharmacists were confident in a technician’s ability to competently prepare vaccines (63.7% vs 91.8%, p <0.001). A statistically significant association was observed between an efficient process for immunizing patients and the likelihood of technician vaccination errors (χ^2^ = 14.36; p <0.01).

**Conclusion:** Pharmacy technicians continue to be a vital part of the healthcare team. Based on survey results, pharmacists and pharmacy technicians responded that technicians are not only competent enough to give immunizations but, should participate in more patient care duties. Multiple states are enacting legislation to include technician vaccine administration as a permanent component of their scope of practice.

**Summary Bullets:** *What is already known about this subject?:* The Public Readiness and Emergency Preparedness (PREP) Act was amended during the COVID-19 pandemic to authorize pharmacy technicians to administer the Advisory Committee on Immunization Practices (ACIP) recommended immunizations and COVID-19 vaccines under pharmacist order. Prior to the COVID-19 pandemic, multiple studies showed that pharmacists recognized pharmacy technicians as vital members of the healthcare team that enhanced the immunization process. In addition, studies found that most pharmacists were comfortable with pharmacy technicians collecting paperwork, processing, and billing vaccinations but did not agree with the idea of the pharmacy technician scope of practice expanding to include immunization administration.

*What does this study add?:* This study evaluates and compares pharmacists’ and pharmacy technicians’ attitudes and experiences associated with technician administered vaccines after the PREP Act authorized technicians to administer vaccines during the COVID-19 pandemic.

**Disclosures:** The authors of this study have no possible financial or personal relationships with commercial entities to disclose that may have a direct or indirect interest in the matter of this study.

**Funding:** None

## INTRODUCTION

On March 11, 2020, the World Health Organization (WHO) declared a global SARS-CoV-2 (COVID-19) pandemic.^1^ The novel COVID-19 virus had a dramatic impact on the American healthcare system in 2020 as evidenced by a 9.7% growth in national healthcare spending and demand.^1^ There was a drastic increase in the utilization of hospital and clinical services for COVID-19 relief.^1^ As a result, elective procedures, general wellness visits, and immunization administration plummeted.^1^ In response to the increased demand for healthcare services, the Department of Health and Human Services (HHS) amended the Public Readiness and Emergency Preparedness (PREP) Act to authorize pharmacy technicians, who are not otherwise authorized in their state, to administer the Advisory Committee on Immunization Practices (ACIP) recommended immunizations and COVID-19 vaccines under pharmacist order during the state of emergency.^1,2^

Idaho became the first state to expand the technician scope of practice to include immunization administration in 2017 with Rhode Island and Utah following shortly after.^3-5^ Other states have been slow to adopt technician administered vaccinations into official law as of late 2019.^3,4^ With pharmacies being the most accessible healthcare destination for the public, it is anticipated that the number of vaccination services given by local pharmacies will continue to increase.^4^ In the future, more states may incorporate pharmacy technician vaccine administration into law as a gateway to increase vaccination rates and improve access to healthcare.^3^

There is limited literature evaluating the overall attitudes and experiences regarding the potential expansion of the pharmacy technician scope of practice to include more patient care duties such as vaccine administration. A cross-sectional study completed by Ohio Northern University in 2014 found that 93% of pharmacists agreed that pharmacy technicians play a vital role in making the immunization process run smoothly, however, the study failed to evaluate the opinions regarding pharmacy technician immunization administration.^4^ Another cross-sectional study conducted in 2018 by the University of Alabama Birmingham found that most pharmacists agreed that pharmacy technicians could competently collect paperwork, process, and bill vaccines but only 24% of pharmacists agreed with the idea of technicians administering vaccine doses.^6^

When it came to pharmacy technician opinion regarding the willingness to perform emerging tasks in the community setting, a cross-sectional study completed in 2018 found that pharmacy technicians showed a moderate to high willingness to perform most emerging tasks except for vaccine administration and fingerstick blood draw which showed a low willingness.^7^ The COVID-19 pandemic started after these studies were conducted and technicians have since been authorized by the PREP Act to administer vaccines under the order of a pharmacist which may have changed the overall attitudes and opinions regarding the expansion of the pharmacy technician scope of practice to include vaccine administration.^1,2^ The primary objective of this cross-sectional study is to evaluate and compare the attitudes and opinions associated with technician administered immunizations from pharmacists and pharmacy technicians in the community setting.

## METHODS

### Study Design & Implementation

This is a cross-sectional study approved by the University of New England IRB committee evaluating the attitudes and experiences of pharmacists and pharmacy technicians regarding technician administered vaccines. Participants in both groups had to be at least 18-years of age and complete a survey consent form. Pharmacy technicians were selected to complete the survey at a grocery chain pharmacy located across 5 states in the Northeast if they completed the 20-hour American Pharmacist Association (APhA) immunization certification program and administered at least 1 immunization throughout the course of the pandemic. Their supervising pharmacists were also selected to complete a similar survey.

The survey was drafted using Microsoft Forms. Two versions of the survey were created to evaluate the primary endpoint, one version was for supervising pharmacists to complete, and the other version for immunization certified pharmacy technicians to complete. Both versions of the survey consisted of a 20-item peer-reviewed questionnaire with 7-items inquiring about background information. To ensure that the survey responses remained anonymous, no identifiable information was collected including employee identification number, email, name, address, or phone number. Participants were asked to rate their level of agreement with each statement on a scale from 1 to 7 with 1 being strongly disagree, 4 being neutral, and 7 being strongly agree. Responses for each survey question that were rated 1-7 were further aggregated into agree, neutral, and disagree for statistical analysis.

### Participant Recruitment

Supervising pharmacists and their immunization certified technicians were informed about the cross-sectional study analyzing the attitudes and experiences surrounding technician administered vaccines through a corporate wide pharmacy email. The email explained the purpose of the study and how the survey results would be managed as well as where to find the survey and survey consent form online if one decided to participate. The surveys were launched on February 1, 2022, and participants had 14 days to complete them. A reminder email was sent 6, 8, and 10 days after the survey was launched to help improve participation.

### Statistical Analysis

Descriptive statistics were calculated to examine pharmacist and pharmacy technician beliefs regarding technician administered vaccination by uploading the results from Microsoft Forms into Microsoft Excel. Microsoft Excel was used to organize and compare the survey responses into tables and charts. Chi square tests (χ^2^) were utilized to evaluate the correlation between responses.

## RESULTS

### Respondent Characteristics

Supervising pharmacists and immunizing pharmacy technicians in a grocery chain pharmacy across 6 states in the Northeast were notified about the study and how to participate through a corporate wide email. Participants were given 14 days to complete the survey. A total of 268 survey responses were obtained with 171 from pharmacists (63.8%) and 97 from technicians (36.2%).

The survey population was largely female (69%) and most were practicing in Maine (36.6%) or New York (29.1%). Respondents were well distributed across age and pharmacy experience overall (Table 1). However, supervising pharmacists were more likely to be male, older, and more experienced in pharmacy when compared to immunizing technicians (p<0.05).

**Table 1.** Survey Respondent Characteristics

|  | Characteristic | Pharmacist<br>n(%) | Technician<br>n(%) | Total<br>n(%) |
| --- | --- | --- | --- | --- |
| Gender* | Male | 56 (32.7) | 12 (12.4) | 68 (25.4) |
|  | Female | 105 (61.4) | 80 (82.5) | 185 (69.0) |
|  | Non-binary | 0 (0.0) | 2 (2.1) | 2 (0.8) |
|  | Unknown | 10 (5.8) | 3 (3.1) | 13 (4.9) |
| Age (years)* | 18-30 | 34 (19.9) | 39 (40.2) | 73 (27.2) |
|  | 31-40 | 64 (37.4) | 24 (24.7) | 88 (32.8) |
|  | 41-50 | 38 (22.2) | 17 (17.5) | 55 (20.5) |
|  | 51-60 | 29 (17.0) | 14 (14.4) | 43 (16.0) |
|  | 60+ | 6 (3.5) | 3 (3.1) | 9 (3.4) |
| Practicing State | Maine | 58 (33.9) | 40 (41.2) | 98 (36.6) |
|  | Massachusetts | 9 (5.3) | 9 (9.3) | 18 (6.7) |
|  | New Hampshire | 24 (14.0) | 15 (15.5) | 39 (14.6) |
|  | New York | 54 (31.6) | 24 (24.7) | 78 (29.1) |
|  | Vermont | 10 (5.8) | 7 (7.2) | 17 (6.3) |
|  | Multiple States | 16 (9.4) | 2 (2.1) | 18 (6.7) |
| Work Experience (years)* | 0-5 | 39 (22.8) | 36 (37.1) | 75 (28.0) |
|  | 6-10 | 30 (17.5) | 30 (30.9) | 60 (22.4) |
|  | 11-20 | 50 (29.2) | 21 (21.6) | 71 (26.5) |
|  | 20+ | 52 (30.4) | 10 (10.3) | 62 (23.1) |
| Total |  | 171 (100) | 97 (100) | 268 (100) |

Supervising pharmacists were more likely to be male, older, and more experienced in pharmacy when compared to immunizing technicians ( $p < 0.05$ )

### Pharmacist Attitudes & Experiences

A total of 171 supervising pharmacists responded to the survey and underwent review as shown in Table 2. The majority of supervising pharmacists agreed that pharmacy technician administration improved the workflow of immunization services in the pharmacy (76.6%) and that technicians could safely administer vaccines (87.1%). Pharmacists also agreed that technicians could competently process and bill for vaccine services (90.6%). In addition, pharmacists reported that technician administered vaccines helped them focus on pharmacist related duties (73.7%) and that most patients were comfortable with technicians administering their immunizations (80.1%). Approximately 56.1% of supervising pharmacists responded that the technician scope of practice should expand to include more patient care duties such as immunization and point of care testing. Lastly, most pharmacists agreed that their pharmacy had an efficient process for immunizing patients (87.7%), but only 58.5% of pharmacists agreed that they had sufficient space to immunize patients.

**Table 2.** Supervising Pharmacist Survey Response Result Summary (n=171)

| Survey Statement | Supervising Pharmacist Response |  |  |
| --- | --- | --- | --- |
|  | Agree n(%) | Neutral n(%) | Disagree n(%) |
| Technicians who have been trained to administer vaccines have improved the workflow of vaccine services in the pharmacy | 131 (76.6) | 27 (15.8) | 13 (7.6) |
| Pharmacy technicians safely administer vaccines | 149 (87.1) | 14 (8.2) | 8 (4.7) |
| Pharmacy technicians competently prepare vaccines for administration | 109 (63.7) | 41 (24.0) | 21 (12.3) |
| Pharmacy technicians competently process vaccine prescriptions including billing | 155 (90.6) | 4 (2.3) | 12 (7.1) |
| Technician vaccine administration has increased my ability to focus on my duties as a pharmacist | 126 (73.7) | 20 (11.7) | 25 (14.6) |
| It is challenging to supervise technician vaccine administration while performing my other duties as a pharmacist | 73 (42.7) | 28 (16.4) | 70 (40.9) |
| Technician vaccine administration increases the likelihood of vaccination errors | 37 (21.6) | 38 (22.3) | 96 (56.1) |
| The technician scope of practice should expand to include more patient care duties under the supervision of a pharmacist | 96 (56.1) | 38 (22.3) | 37 (21.6) |
| Only technicians who have the CPhT credential should be trained to administer immunizations | 137 (80.1) | 13 (7.6) | 21 (12.3) |
| Most patients appear to be comfortable with vaccines administered by a pharmacy technician | 187 (80.1) | 29 (17.0) | 5 (2.9) |
| Vaccine administration training should be required for technicians who practice in community pharmacy settings | 66 (38.6) | 34 (19.9) | 71 (41.5) |
| The pharmacy where I work has an efficient process for immunizing patients | 150 (87.7) | 14 (8.2) | 7 (4.1) |
| The pharmacy where I work has sufficient space to immunize patients | 100 (58.5) | 21 (12.3) | 50 (29.2) |

Responses for each survey question that were rated from 1-7 (1=strongly disagree, 4=neutral, 7=strongly agree) were further aggregated into agree, neutral, and disagree

There were some challenges indicated by pharmacists’ survey responses. Many agreed that it was challenging to supervise technician administered immunizations while performing other duties as a pharmacist (42.7%). In addition, significantly fewer pharmacist survey responses agreed that technicians could competently prepare vaccines when compared to technician responses (63.7% vs 92%; p<0.001). A total of 21.6% of pharmacists and 10.3% of technicians responded that technician administered vaccines increased the likelihood of vaccine related errors. Furthermore, as shown in Table 4, there was a significant association observed between an efficient process for immunizing patients and the likelihood of technician vaccination errors (χ^2^ = 14.36; p <0.01).

### Pharmacy Technician Attitudes & Experiences

A total of 97 immunizing pharmacy technicians responded to the survey and underwent review as shown in Table 3. Most of the pharmacy technician responses were similar to their supervising pharmacists. Most pharmacy technicians agreed that technician administered immunizations improved the workflow of immunization services in the pharmacy (82.5%). Immunizing technicians also agreed that they could competently process and bill vaccine services (99%), competently prepare vaccines for administration (91.8%), and safely administer vaccines to patients (96.9%) without increasing the likelihood of vaccine errors (78.3%). Approximately 69% of technicians agreed that the pharmacy technician scope of practice should expand to include more patient care duties such as immunizations and point of care testing. In addition, many responded that patients were comfortable with a technician administering vaccines (86.6%) and that their supervising pharmacists were accessible for questions about immunizations and safety (96.9%). Over half of technician responses showed that administering vaccines did not prevent them from doing other technician duties effectively such as filling, data entry, and third party (51.5%). Lastly, technicians reported that their pharmacy had an efficient process for vaccinating patients (86.6%) and had sufficient space for vaccine administration (67%).

**Table 3.**
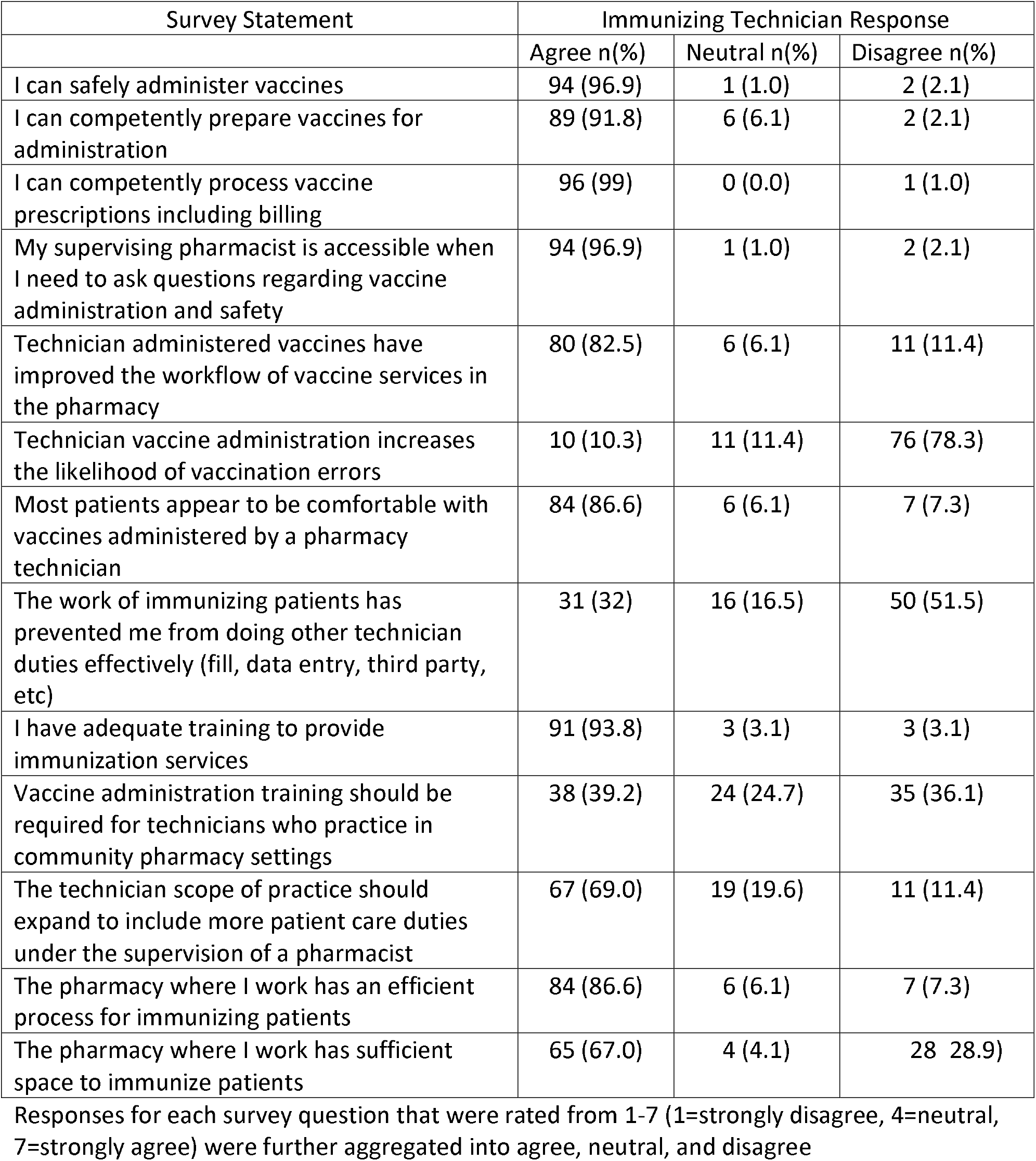
Immunizing Pharmacy Technician Survey Response Summary (n=97)

**Table 4.**
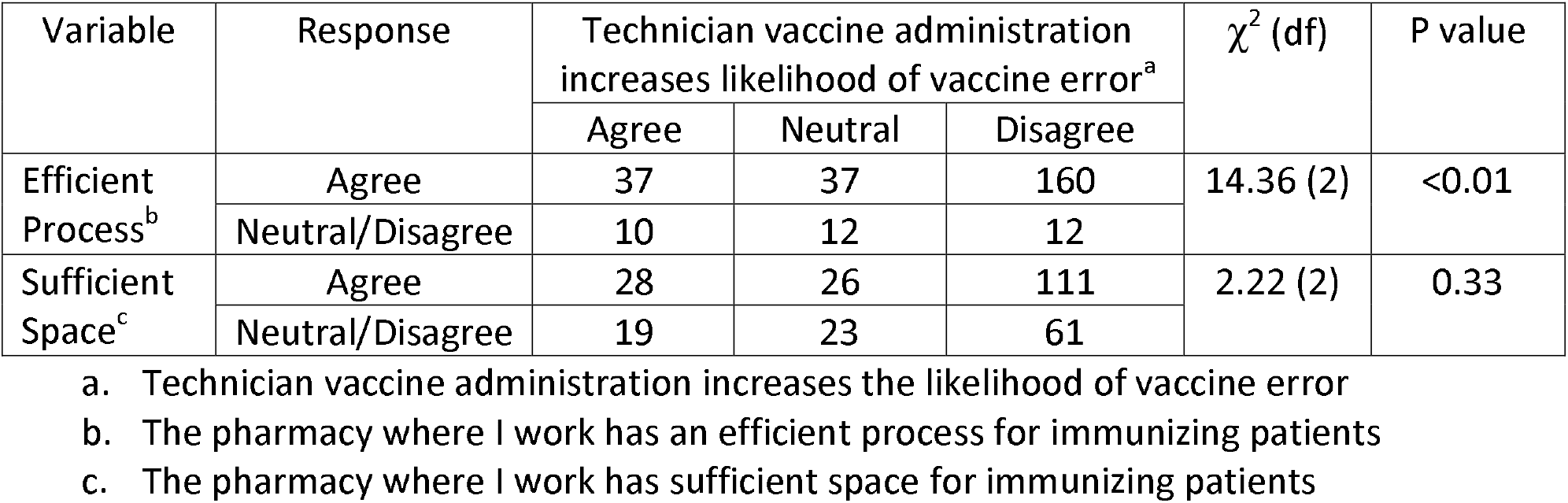
Cross tabulation showing observed cell counts between pharmacist and pharmacy technician perception of an efficient process or sufficient space for immunization services and the likelihood of vaccination errors associated with technician vaccine administration

## DISCUSSION

This cross-sectional study aimed to evaluate and compare the attitudes and experiences of supervising pharmacists and immunizing technicians regarding technician administered vaccines through anonymous, peer-reviewed surveys. There is limited literature reporting the attitudes and experiences of technician administered vaccines among pharmacists and technicians. Prior studies have shown that pharmacist survey responses agreed that pharmacy technicians improve the immunization process but none of the studies had an in-depth evaluation of pharmacist and pharmacy technician opinion regarding the expansion of the technician scope of practice to include vaccine administration.^4,6,7^ These studies were prior to the PREP Act which authorized technicians to give vaccines under pharmacist order and consequently there’s a need to re-evaluate the attitudes, experiences, and challenges associated with technician administered vaccines in the context of the pandemic.^2^

Adult immunization rates for pneumococcal, influenza and herpes zoster vaccination did not reach public health targets before 2020 and the COVID-19 pandemic worsened immunization rates.^1,4^ Local pharmacies are one of the most accessible healthcare services and have been vaccinating the public for years with success.^4,8^ The demand for vaccine services has increased among pharmacies during the pandemic and it is anticipated that the demand for vaccine services will continue to increase.^1,2^ The PREP Act temporarily authorized pharmacy technicians to administer ACIP recommended immunizations and COVID-19 vaccines under pharmacist order during the state of emergency.^2^ Prior to the pandemic, Adams et al insightfully called for regulatory changes to state policy in order to enable technicians to administer vaccines.^5^ Multiple states are now enacting legislation to include technician vaccine administration as a permanent component of their scope of practice.^3^

This study found that most pharmacists and pharmacy technicians responded that technicians could safely administer vaccines (87.1% and 96.9%, respectively) and competently process and bill vaccine services (90.6% and 99.0%, respectively). In addition, both participant populations responded that technician administered vaccines improved the workflow of vaccine services (76.6% and 82.5%, respectively) without increasing the likelihood of vaccine errors (56.1% and 78.3%, respectively). When compared with technicians, fewer pharmacists were confident in a technician’s ability to competently prepare vaccines (63.7% vs 91.8%, p <0.001). A statistically significant association was observed between an efficient process for immunizing patients and the likelihood of technician vaccination errors (χ^2^ = 14.36; p <0.01). Furthermore, approximately 56.1% of pharmacists and 69.0% of technicians agreed that the technician scope of practice should expand to include more patient care duties under pharmacist supervision.

### LIMITATIONS

This is a cross-sectional study that evaluated the primary endpoint through the completion of surveys in which non-response bias is of concern. Furthermore, respondents could have interpreted the statements differently which may have impacted the results. A convenience sample from a grocery pharmacy chain located in 6 states across the Northeast was utilized which limits the generalizability of the results.

## CONCLUSION

Pharmacy technicians continue to play a vital role on the healthcare team. Most supervising pharmacists and immunizing technicians surveyed support the recent expansion of the technician scope of practice to include the administration of immunizations. Furthermore, both group’s survey responses favor the expansion of the pharmacy technician scope of practice to include more patient care duties under the supervision of the pharmacist. Further studies are needed in order to determine trends amongst pharmacist and technician attitudes and experiences in the United States.

## Data Availability

All data produced in the present study are available upon reasonable request to the authors

## REFERENCES

1. Office of the Actuary. National Health Spending in 2020 Increases Due to Impact of COVID-19 Pandemic. CMS. 2021 Dec 15. Accessed 2022 May 5. Available at: https://www.cms.gov/newsroom/press-releases/national-health-spending-2020-increases-due-impact-covid-19-pandemic

2. Office of the Assistant Secretary of Health. Guidance for PREP Act Coverage for Qualified Pharmacy Technicians and State-Authorized Pharmacy Interns for Childhood Vaccines, COVID-19 Vaccines, and COVID-19 Testing. HHS. 2020 Oct 20. Accessed 2022 May 5. Available at: https://www.hhs.gov/sites/default/files/prep-act-guidance.pdf

3. Eid D, Osborne J, Borowicz B. Moving the Needle: A 50-State and District of Columbia Landscape Review of Laws Regarding Pharmacy Technician Vaccine Administration. Pharmacy (Basel). 2019;7(4):168. Published 2019 Dec 10. doi:10.3390/pharmacy7040168

4. Aldrich S, Sullivan D. Assessing Pharmacists’ Attitudes and Barriers Involved with Immunizations. Innov Pharm. 2014;5(2):1. Published 2014 Jan 1. doi:10.42926/iip.v5i2.336

5. Adams AJ, Desselle SP, McKeirnan KC. Pharmacy Technician-Administered Vaccines: On Perceptions and Practice Reality. Pharmacy (Basel). 2018;6(4):124.

6. Kulczycki A, Grubbs J, Hogue MD, Shewchuk R. Community chain pharmacists’ perceptions of increased technicians’ involvement in the immunization process. J Am Pharm Assoc (2003). 2021;61(5):596–604. doi:10.1016/j.japh.2021.04.017

7. Doucette WR, Schommer JC. Pharmacy Technicians’ Willingness to Perform Emerging Tasks in Community Practice. Pharmacy (Basel). 2018;6(4):113. Published 2018 Oct 12. doi:10.3390/pharmacy6040113

8. Westrick SC, Patterson BJ, Kader MS, Rashid S, Buck PO, Rothholz MC. National survey of pharmacy-based immunization services. Vaccine. 2018;36(37):5657–5664.

